# Radiomic-Based Prediction of Lesion-Specific Systemic Treatment Response in Metastatic Disease

**DOI:** 10.1101/2023.09.22.23294942

**Authors:** Caryn Geady, Farnoosh Abbas-Aghababazadeh, Andres Kohan, Scott Schuetze, David Shultz, Benjamin Haibe-Kains

## Abstract

Despite sharing the same histologic classification, individual tumors in multi metastatic patients may present with different characteristics and varying sensitivities to anticancer therapies. In this study, we investigate the utility of radiomic biomarkers for prediction of lesion-specific treatment resistance in multi metastatic leiomyosarcoma patients. Using a dataset of n=202 lung metastases (LM) from n=80 patients with 1648 pre-treatment computed tomography (CT) radiomics features and LM progression determined from follow-up CT, we developed a radiomic model to predict the progression of each lesion. Repeat experiments assessed the relative predictive performance across LM volume groups. Lesion-specific radiomic models indicate up to a 4.5-fold increase in predictive capacity compared with a no-skill classifier, with an area under the precision-recall curve of 0.70 for the most precise model (FDR = 0.05). Precision varied by administered drug and LM volume. The effect of LM volume was controlled by removing radiomic features at a volume-correlation coefficient threshold of 0.20. Predicting lesion-specific responses using radiomic features represents a novel strategy by which to assess treatment response that acknowledges biological diversity within metastatic subclones, which could facilitate management strategies involving selective ablation of resistant clones in the setting of systemic therapy.

**Highlights:**

- Intensity values in CT scans and their corresponding spatial distribution convey important information.
- A model to predict lesion-specific response to systemic treatment using image-derived features is proposed.
- Up to a 4.5-fold increase in predictive capacity compared to a no-skill classifier was obtained, with AUPRC of 0.70 for the most precise model (FDR = 0.05).
- Assessing treatment response on a lesion-level acknowledges biological diversity within metastatic subclones, which could facilitate management strategies involving selective ablation of resistant clones in the setting of systemic therapy.

## Introduction

Cancer is a dynamic disease characterized by the development of rapidly-dividing abnormal cells and is a leading cause of death worldwide [1]. As cancer develops, subpopulations of cells emerge with distinct genotypes and phenotypes, harboring divergent biological behaviors [2]. The net result is increased cancer heterogeneity over time: individual patients, lesions, and cell populations with varying sensitivities to anticancer therapies [3]. Cancer heterogeneity as such is associated with inferior clinical outcomes. In this study we focus on inter-metastatic heterogeneity, which refers to the heterogeneity among different metastatic lesions of the same primary tumor. Widespread metastases are the primary cause of death in cancer patients [4]. The vast majority of patients with solid tumors die because of metastasis to the liver, brain, lung, or bone. Patients who relapse with a single metastatic lesion can occasionally be cured by surgery or radiotherapy, but single metastases are the exception rather than the rule [5], [6]. Unfortunately, metastatic sites develop unique phenotypes and genotypes [7], [8]. As such, eradicating a subset of metastatic lesions in a patient is not likely to provide adequate long-term disease control. From that perspective, identifying treatment resistant tumors may facilitate combination therapies to provide a more successful treatment outcome [5].

Radiological imaging has a critical role in cancer diagnosis and in evaluating treatment resistant tumors. In particular, radiomics has garnered much attention from the research community for its potential predictive power for treatment outcomes and cancer genetics [9], [10]. Radiomics has become an active field of research, allowing scientists to extract quantitative features from readily-available radiological images and assess their potential as non-invasive biomarkers [9], [10]. These features can provide information about intensity, shape, volume, and texture of tumor phenotypes [11]–[14]. In this study, we investigate the utility of radiomic-based biomarkers for prediction of lesion-level systemic treatment response, which could help to get a more comprehensive view on the overall patient status. This type of response prediction has been recently initiated within the context of imaging studies. Correlation between computed tomography (CT) textural features with pathological features and clinical outcome has been demonstrated in liver metastases [15], [16]. More recently, lesion level immunotherapy response prediction using image-derived or radiomic features has been explored across a range of metastatic sites with promising results [17], [18]. To the best of the authors’ knowledge however, there hasn’t yet been a successful implementation of a predictive model that can accurately predict the response of individual lesions to standard systemic treatments for metastatic patients.

Soft tissue sarcomas (STSs) are cancers of connective and supportive tissues in the body; they are rare, heterogeneous, and notoriously difficult to manage clinically. Patients presenting with locally advanced or metastatic disease have dramatically lower rates of survival than those with non-advanced localized disease [19]–[21]. In particular, leiomyosarcoma (LMS), one of the more common STS subtypes, tend to be biologically aggressive tumors with high metastatic potential and local recurrence rates; as a result, systemic therapy plays an important role in the multimodality treatment strategy [22]. Radiologic assessment of therapeutic efficacy then becomes a crucial task so that ineffective therapies can be switched out for alternative and potentially more active regimens. Radiomic features are particularly well-suited to interrogating LMS, where the spatial clustering of enhancing and nonenhancing voxels map histologically to viable and necrotic tumor components [22], [23]. Identifying radiomic biomarkers of treatment response could help to identify LMS patients who could benefit from alternative and potentially more active regimens towards improved outcomes. To this end, we analyzed all visible pulmonary lesions to evaluate the predictive value of CT-derived radiomic biomarkers in metastatic LMS receiving cytotoxic chemotherapy.

## Methods

### Data Collection and Generation Participants

Our patient cohort included those patients who participated in a randomized Phase III, multicenter, open label study comparing Doxorubicin Monotherapy (DM) versus Doxorubicin plus Evofosfamide (DE) in locally advanced, unresectable or metastatic soft-tissue sarcoma (TH CR-406/SARC021, NCT01440088). Full trial protocol and results were published by Tap et. al.[24]. A total of 640 patients were enrolled; the primary endpoint of the trial was overall survival. Contrast-enhanced CT obtained prior to treatment and after 2 cycles of systemic therapy were available for analysis in 180 leiomyosarcoma (LMS) patients.

### Image Segmentation

A database of serial CT imaging was obtained from the Sarcoma Alliance for Research through Collaboration (SARC). Chest CT images at two time points (baseline/prior to treatment and after 2 cycles of systemic therapy/at follow-up) were uploaded into the open-source software 3D Slicer (https://www.slicer.org/). All lung lesions which were identifiable on CT were segmented and subsequently reviewed by a radiologist with 10 years of experience. Lesions were considered identifiable if they were measurable at baseline as defined in RECIST 1.1 (minimum diameter of 10mm in the longest plane of measurement [25]) and confidently located at the second time point. Further, lesions that could not be accurately discriminated from surrounding tissues (ie, lung nodule adjacent or within atelectasis) or from other adjacent lesions at baseline or follow-up CTs (i.e., confluent metastases) were not delineated and excluded.

### Morphological Image Processing

Imaging data and lesion segmentations were resampled to a common 1 × 1 × 1 mm^3^ voxel size using bi-linear and nearest neighbor interpolation, respectively. The original lesion segmentation denoting the lesion boundaries was analyzed as the intratumoral region (whole lesion). From this intratumoral mask, an additional three segmentations containing sub-volumes of interest were generated using standard morphological processing (**Figure 1**): the lesion core and internal and external rims. A ball-shaped structuring element, with a radius spatially equivalent to 2 mm, was used to perform the morphological operations.

**Figure 1:**
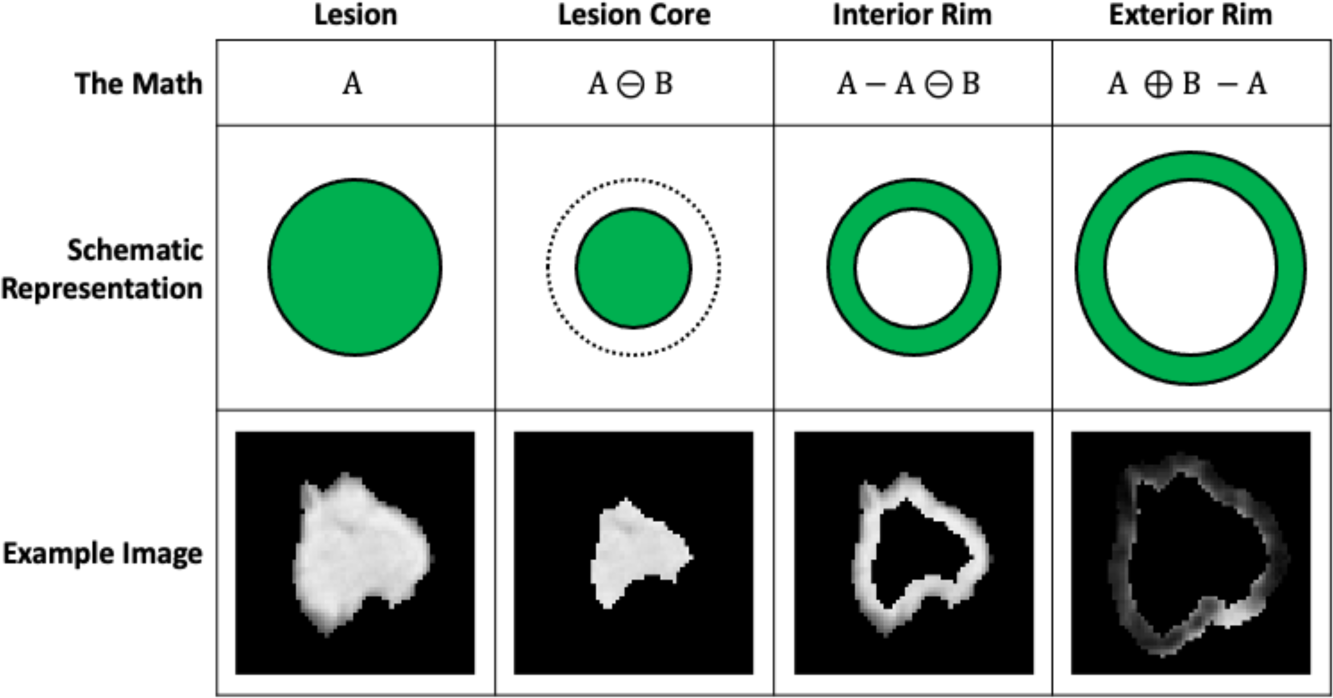
Standard morphological processing to define sub-volumes of interest. Corresponding schematic representations and examples are shown for illustrative purposes.

The intratumoral mask was eroded to isotropically contract the lesion boundary; the remaining voxels in the mask were analyzed as the lesion core. Voxels in the lesion core were eliminated from the intratumoral mask, leaving a mask containing a ring of lesion analyzed as the internal peritumoral region or internal rim. The intratumoral mask was dilated to isotropically expand the lesion boundary. Voxels included in the original intratumoral mask were eliminated from the dilated mask, leaving a mask containing a ring of lung parenchyma analyzed as the external peritumoral region or external rim.

### Radiomic Feature Extraction

Radiomic features were extracted per the process championed by the Imaging Biomarker Standardization Initiative (IBSI) [13], which is implemented with the open-source software package PyRadiomics (version 3.0.1) [12]. For each lesion and corresponding sub-volumes of interest, a total of 1648 features were calculated; features include first-order statistical measures, histogram, shape and size descriptions, Grey Level Co-occurrence Matrix (GLCM), Grey Level Run Length Matrix (GLRLM), Grey Level Size Zone Matrix (GLSZM) and Neighboring Grey Tone Difference Matrix (NGTDM) features.

### Data Analysis

#### Response Categorization

A lesion-wise evaluation of relative change in volume between baseline and follow-up was performed using PyRadiomics (version 3.0.1); the shape descriptive radiomic feature ‘VoxelVolume’ of the intratumoral mask defined the lesion volume at baseline and after 2 cycles of systemic therapy, respectively. Population-specific outliers were removed using a median absolute deviation outlier detection method, based on the lesion volume at baseline [26]. Percent change in volume with respect to baseline was evaluated as a metric of treatment response. A response threshold, T_r_, was defined such that lesions that experienced a positive volume change greater than T_r_ were labeled as ‘progressive’; all other lesions were labeled as ‘non-progressive’.

#### Response Prediction

Firstly, features not robust with respect to the segmentation process were removed as previously described [27]. Briefly, each lesion segmentation was eroded and dilated isotropically by 1 mm (implemented with scikit-image module, version 1.6.2) and radiomic features that exhibited a concordance correlation coefficient below 0.8 between the original segmentation and each of the derived ones were dropped [28]. Then, a correlation analysis was performed: radiomic features were hierarchically clustered using the agglomerative unweighted pair group method with arithmetic mean (implemented with scipy version 1.6.2). A distance metric of 0.5 was used to define the main clusters; from these clusters, the medoid of each cluster was selected as a representative feature. To eliminate potential confounding effects with lesion volume, correlation was assessed and cluster-generated features with an absolute volume-correlation coefficient greater than 0.20 were removed [29].

From the reduced feature set, up to 4 radiomic features were selected for model inclusion using the SelectKBest method from the sklearn module (version 1.1.1) [30]. For classification tasks, this supervised method calculates the ANOVA F-values between the target and each feature, sorts them and selects the K best features. Logistic modeling was considered for the classification task as a baseline model due to its robustness, interpretability, and generalizability, particularly due to the limited size of the dataset (a common feature of datasets related to rare diseases like LMS). The model architecture was fine-tuned using the GridSearchCV method from the sklearn module (version 1.1.1); details regarding this process can be found in Supplementary. Five models were tested in total for each arm: one model with baseline volume only, one model with baseline volume plus one selected radiomic feature, and so on. To account for any potential class imbalance between response categories, we employed a stratified 10-fold cross validation to fit the data for each modeling strategy using sklearn’s RepeatedStratifiedKFold [31]. This procedure splits the dataset in such a way that preserves the same class distribution (i.e., the same percentage of sample of each class, in our case progressive and non-progressive lesions) in each subset/fold as in the original dataset. However, a single run of StratifiedKFold might result in a noisy estimate of the model’s performance, as different splits of the data might result in very different results. As its name suggests, RepeatedStratifiedKFold allows improving the estimated performance of a machine learning model, by simply repeating the cross-validation procedure multiple times (according to the n_repeats value, which was set to 5 for this study), and reporting the mean result across all folds from all runs. This mean result is expected to be a more accurate estimate of the model’s performance. The best performing model was defined as the model which minimized the negative log likelihood. Model performance was quantified using Area Under the Curve for the Receiver Operating Characteristic (AUROC) and Precision-Recall (AUPRC) curves as well as the evaluation of Matthew’s Correlation Coefficient (MCC) [32]–[34]. A paired Wilcox test was used to test for differences in performance distribution for the model with volume alone versus the best performing model.

#### Sensitivity Analysis

The response prediction task was repeated for an additional two subgroups. A median absolute deviation (MAD) outlier detection method was applied to identify population-specific outliers, based on the lesion volume at baseline [26]. The first subgroup was isolated by removing the population-specific outliers and the second subgroup consisted of the outliers themselves. For the latter case where smaller sample sizes are expectedly smaller, fewer folds were considered for the cross-validation, on the condition that each train/test group of data samples was representative of the broader dataset. A modified Z-score calculated using MAD is considered a robust measure to identify outliers. It replaces standard deviation or variance with median deviation and the mean with the median. The result is a method that isn’t as affected by outliers as using the mean and standard deviation. Lesions were considered outliers if their absolute modified Z-score was greater than 3 [35].

#### Statistics

Descriptions of the patient and lesion population are expressed in terms of median values and the interquartile range (IQR). Relative volume change was assessed as a continuous variable and converted into a binary response category by imposing the response threshold T_r_; frequency and percentage are used to describe the response category distribution. Performance of the predictive models are expressed in terms of their mean with associated 95% confidence interval (CI); significance was evaluated by permutation tests (1000 data permutations) [36]. The False Discovery Rate (FDR) approach was used for multiple testing corrections [37].

## Results

### Data Collection and Generation

A total of 80 patients were found to have at least one lesion that met the inclusion criteria for our study. Of these patients, 33 received Doxorubicin Monotherapy (DM) and 47 received Doxorubicin plus Evofosfamide (DE). The number of contoured lesions per patient ranged from 1-11, with 54/80 or 67.5% of patients with two or more lesions; a total of 202 contoured lesions were included in the analysis, of which 90 received DM and 112 received DE. Median volume of delineated lesions at baseline was 4.17 cc (IQR 2.26 to 9.68). Median time between baseline and follow-up CT was 1.67 months (IQR 1.47 to 1.87).

Radiomic features were extracted from original images as well as from different image transformations including five Laplacian of Gaussian filters (σ = 1.0, 2.0, 3.0, 4.0, 5.0 mm), eight wavelets decompositions, and four non-linearities (exponential, square, square root and logarithm). This generated a set of 6,592 features (1,648 features per sub-volume of interest X 4 sub-volumes of interest). After evaluating robustness to segmentation, 1,452 features (363 features per sub-volume of interest X 4 sub-volumes of interest) were removed. Unsupervised feature selection using clustering identified 75 features representative of highly correlated sets of radiomics features. After removing radiomic features at a volume-correlation coefficient threshold of 0.20, 32 features remained.

### Data Analysis

Using a response threshold (T_r_) of 50%, the fraction of progressive lesions was 15.6% and 24.1% for the DM and DE regimes, respectively (Figure 2). Of the patients with 2 or more lesions, 18.5% exhibited differences in individual lesion response. Significant differences in baseline volume were observed between lesion response categories (Figure 3).

**Figure 2:**
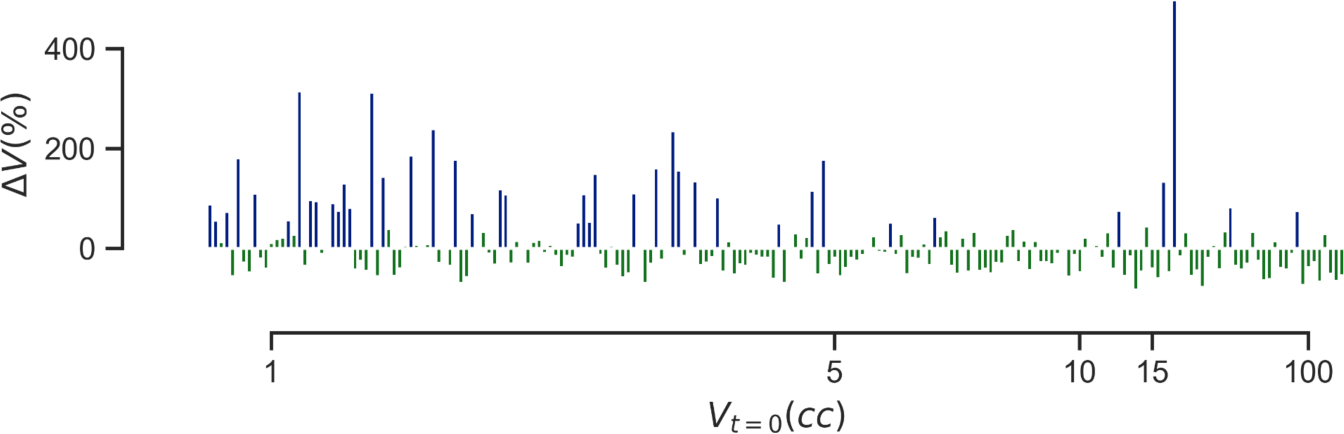
Distribution of relative volume change by lesion volume at baseline. Lesions are colored by response category as determined by T_r_: blue indicates a progressive lesion; green indicates a non-progressive lesion. Lesions are displayed in ascending order of baseline volume.

**Figure 3:**
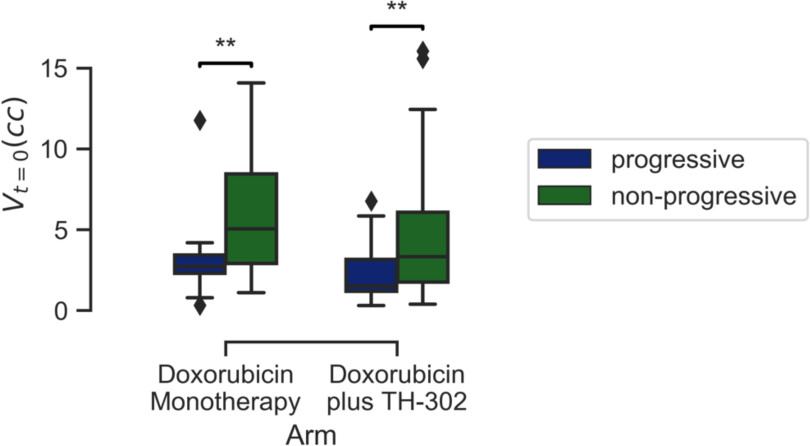
Significant differences observed in lesion volume at baseline by treatment regimen. Population-specific outliers in terms of baseline volume removed for visualization purposes. Annotation for FDR: {‘**’: 0.001 < p <= 0.01}.

For response prediction, 5 models were cross-validated for each treatment regimen (i.e., 1 model with baseline volume, 1 model with baseline volume and 1 radiomic feature, and so on); this process was repeated for 2 additional subgroups based on lesion volume on baseline. The subgroups were determined using the Median Absolute Deviation (MAD) outlier detection method: lesions that had an absolute modified Z-score less than 3 were considered as the first subgroup. Lesions that had an absolute modified Z-score greater than 3 (i.e., outliers in terms of baseline volume) were considered as the second subgroup. Given the distribution of lesion volume included in this study, lesions were considered outliers if their volume at baseline was greater than 16.05 cc.

For the DM regimen, the best performing model consisted of 3 radiomic features, fitted using all lesions, irrespective of baseline volume (**Table 1**). For the DE regimen, the best performing model consisted of 5 radiomic features, fitted using the outlier subgroup, irrespective of baseline volume (**Table 1**). While the model for the DM regimen reached significance (FDR = 0.05), the model for the DE regimen only trended towards significance (FDR = 0.1). Notably, the performance for the both models was significantly different from the volume-only model (p = 2.64×10^−4^ and 4.82×10^−6^ for the DM and DE models, respectively). With the exception of volume, all selected features were from peritumoral regions (i.e., lesion core, interior rim or exterior rim). Generally speaking, the AUROC was higher than the AUPRC, as expected, given the class imbalance. The AUPRC suggests an approximate 4.5 and 5.6-fold increase in predictive capacity compared to a no-skill classifier for the best performing DM and DE models, respectively. The MCC for the DM model indicates a weakly positive relationship between the features and the classification target, whereas the MCC for the DE model indicates a moderate positive relationship.

**Table 1:**
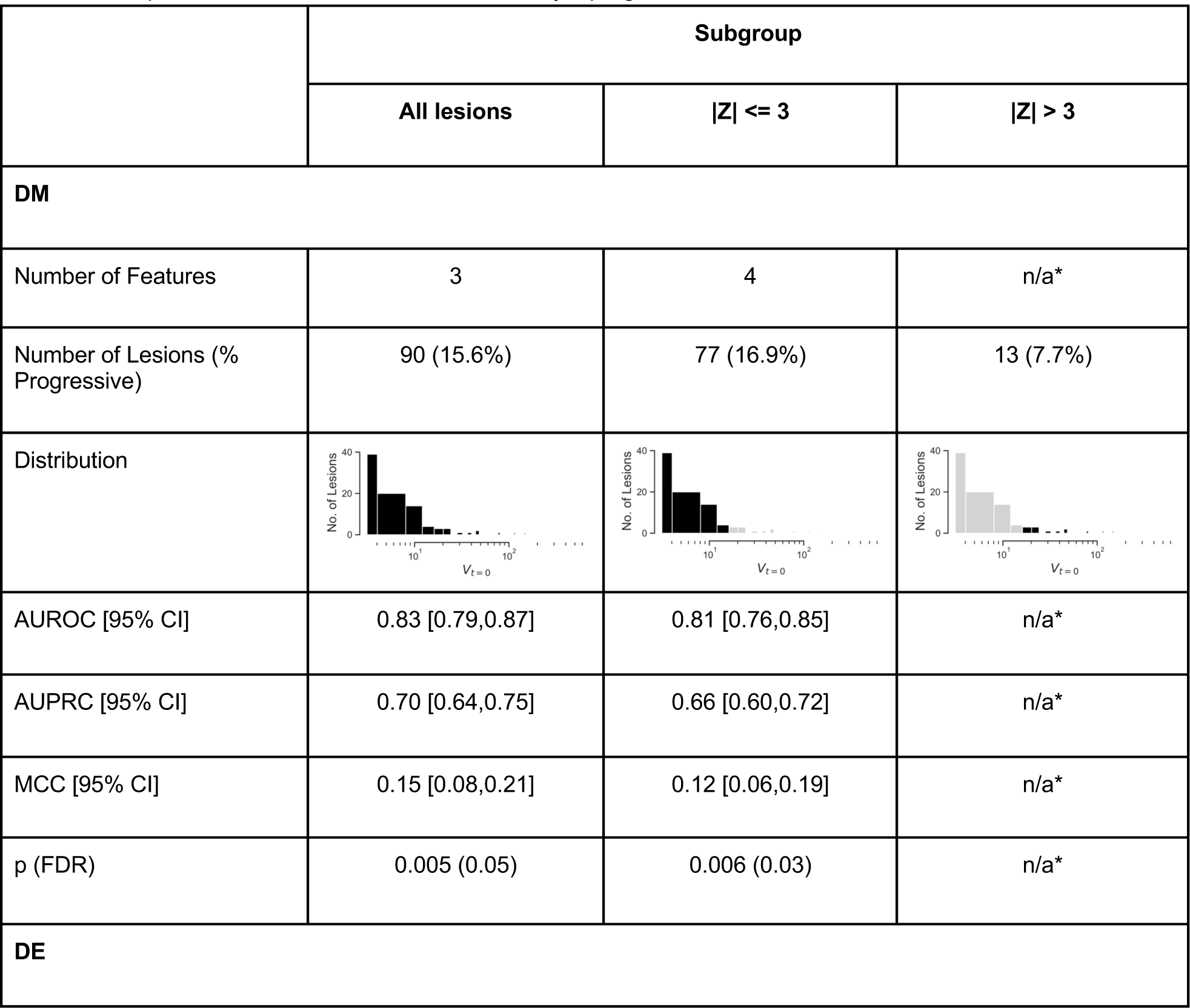
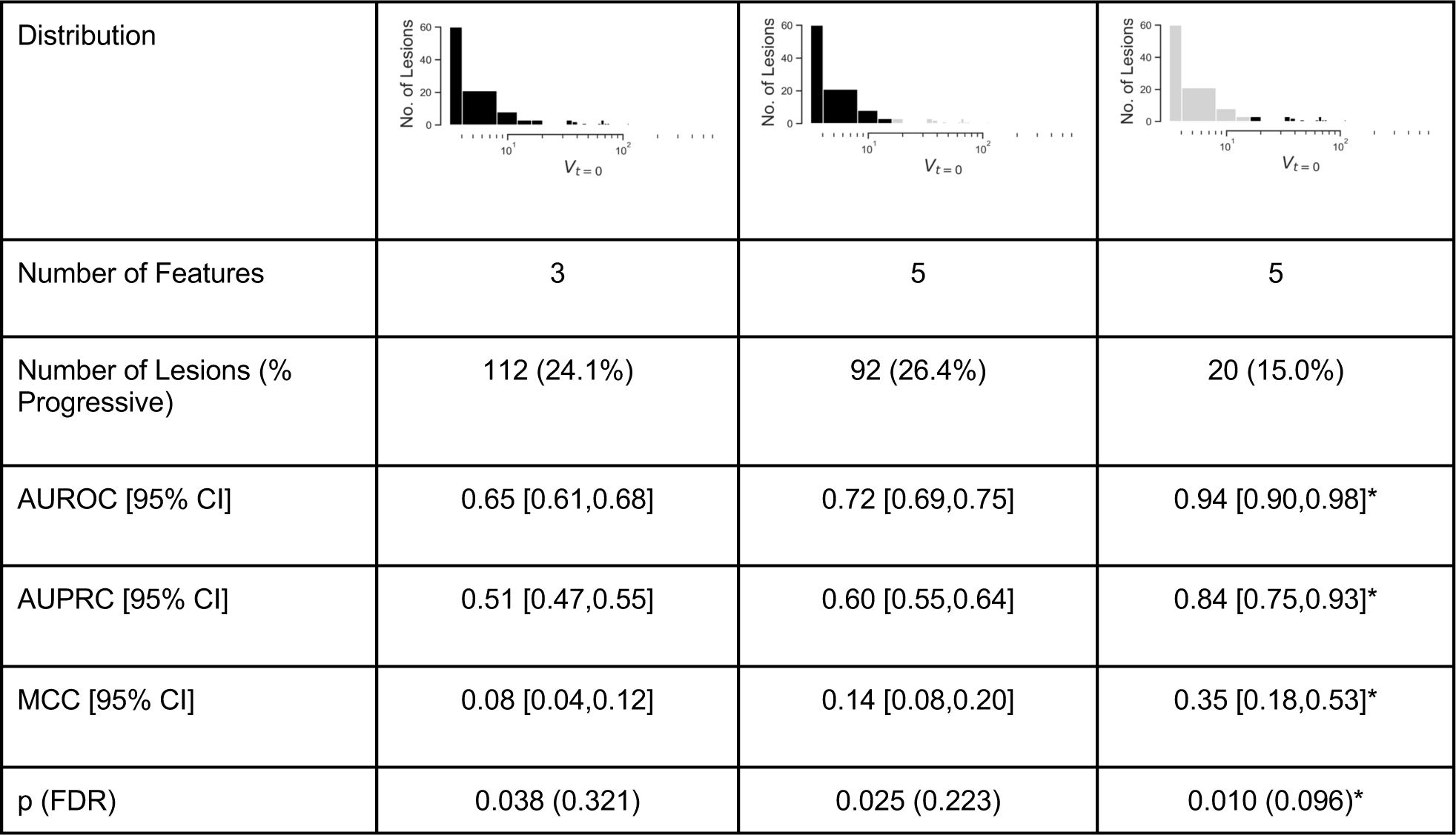
Summary of model performance for the prediction of pulmonary lesion-specific systemic treatment response. Model performance expressed with AUROC, AUPRC and MCC and associated 95% CI; significance indicated by the p-value (FDR) by permutation tests with n=1000 permutations. *For the outlier subgroup receiving DM, the number of progressive lesions was too small (1) to perform cross-validation; for the outlier subgroup of lesions receiving DE, cross-validation was performed with 3-folds as there were only 3 progressive lesions.

|  | Subgroup |  |  |
| --- | --- | --- | --- |
| | All lesions | $\vert Z\vert \leq 3$ | $\vert Z\vert > 3$ |
| **DM** |  |  |  |
| Number of Features | 3 | 4 | n/a* |
| Number of Lesions (% Progressive) | 90 (15.6%) | 77 (16.9%) | 13 (7.7%) |
| Distribution |  |  |  |
| AUROC [95% CI] | 0.83 [0.79,0.87] | 0.81 [0.76,0.85] | n/a* |
| AUPRC [95% CI] | 0.70 [0.64,0.75] | 0.66 [0.60,0.72] | n/a* |
| MCC [95% CI] | 0.15 [0.08,0.21] | 0.12 [0.06,0.19] | n/a* |
| p (FDR) | 0.005 (0.05) | 0.006 (0.03) | n/a* |
| **DE** |  |  |  |
| Distribution |  |  |  |
| Number of Features | 3 | 5 | 5 |
| Number of Lesions (% Progressive) | 112 (24.1%) | 92 (26.4%) | 20 (15.0%) |
| AUROC [95% CI] | 0.65 [0.61,0.68] | 0.72 [0.69,0.75] | 0.94 [0.90,0.98]* |
| AUPRC [95% CI] | 0.51 [0.47,0.55] | 0.60 [0.55,0.64] | 0.84 [0.75,0.93]* |
| MCC [95% CI] | 0.08 [0.04,0.12] | 0.14 [0.08,0.20] | 0.35 [0.18,0.53]* |
| p (FDR) | 0.038 (0.321) | 0.025 (0.223) | 0.010 (0.096)* |

For the outlier lesions receiving DE, there were 20 lesions (of which 3 were progressive); given this distribution, we fitted a model with 3-fold cross-validation instead of 10-fold, such that each split would have at least one progressive lesion. The performance for this model is quite promising; we should however interpret this with some caution due to the sample size and reduced number of folds for the cross-validation.

## Discussion

Monitoring size changes in a cancerous lesion is an important aspect of cancer management, as it can provide valuable information about the tumor’s response to treatment and its potential for progression. Through careful evaluation of these changes in volume over time, healthcare professionals can make informed decisions about treatment strategies, including adjusting treatment doses or switching to alternative therapies [38]. *Predicting* these lesion-specific responses, however, represents a paradigm shift and could present an opportunity to augment existing clinical decision making criteria [39]. In this work, we investigated the utility of radiomic biomarkers for prediction of lesion-specific systemic treatment response in pulmonary metastases and achieved a strong predictive value for lesions receiving doxorubicin monotherapy.

Most published radiomic studies extract radiomic features from only one lesion, even in multi metastatic patients. While analyzing a single lesion may be less time-consuming and involve simpler mathematical models, it may also lead to misevaluation of the cancer inter-lesion heterogeneity [7], [17], [40], [41]. This is especially consequential in the context of predicting patient-specific outcomes such as overall treatment response or patient survival; hence, we sought only to predict tumor response at a lesion level.

Trebeschi *et al* developed a radiomics signature to do exactly this using a cohort of 203 patients receiving immunotherapy (123 patients with non-small cell lung cancer (NSCLC) and 80 melanoma patients) [18]. In their study, although the radiomic biomarker on 303 lesions from the 70 patients of the test reached significant performance (AUROC = 0.66, p < 0.01), only a trend towards significance was obtained from the nodal metastases in melanoma patients (AUROC = 0.64, p = 0.05) and application of the signature was not significant for pulmonary and hepatic melanoma lesions (AUROC = 0.55). However, in their NSCLC cohort, significant performance was observed in pulmonary (AUROC = 0.83, p < 0.001) and nodal metastases (AUROC = 0.78, p < 0.001). Sun *et al* confirmed the association of their previously-validated radiomic score (CD8-Rscore) with response to immunotherapy at lesion level and patient level in 136 patients with advanced melanoma [17]. Notably, predictivity of lesion response using their CD8-Rscore was dependent on the tumor location, with moderate predictive value for subcutaneous lesions (AUROC = 0.65, p = 0.007), and large hepatic (AUROC = 0.70, p = 0.002) and nodal metastases (AUROC = 0.62, p = 0.03), but no significant association was found for pulmonary lesions.

In terms of model performance, our predictive value (AUROC) values align quite closely with these studies. It is important to note, however, that due to the class imbalance in our dataset, AUROC is not the most appropriate measure of model performance. We were most interested in predicting progressive lesions, and the fraction of these were less than 0.5 (i.e., the classes were not balanced). For this, the AUPRC is more informative, as this measure concerns itself with finding all the progressive lesions (recall) without accidentally marking any non-progressive as progressive (precision). The baseline for this metric is equal to the fraction of progressive lesions. For example, the set of lesions receiving DM consists of 15.6% progressive examples and 84.4% non-progressive examples, which sets the baseline AUPRC at 0.156. Obtaining an AUPRC of 0.70 represents a 4.5-fold increase from baseline or a no-skill classifier. This is further reflected in the MCC measure, which takes into account true and false positives and negatives and is generally regarded as a balanced measure. The stark difference observed between the metrics presented here underscore the need for thoughtful choice when it comes to assessing model performance [34].

The aforementioned studies that perform lesion level response prediction align with existing response criteria in that a relative change in *diameter* was evaluated between baseline and follow-up [38]. In our study, we evaluated a relative change in *volume* to define lesion level response where, to our knowledge, no such response criteria currently exist. Results from the literature agree that volume measurement is a method with superior performance in lung tumor sizing, as well as in assessing tumor growth [42], [43]. However, measuring tumor volume is not always straightforward, as tumors can have irregular shapes and may be difficult to accurately measure using imaging techniques [44]. Reported volume measurement errors in lung lesions vary between 20% and 25%; therefore, any relative change in volume beyond this could be considered to define a significant growth [45]–[47]. In our study, we implemented a conservative response threshold of 50%.

Using this threshold, we observed that in patients with 2+ lung metastases, 18.5% of patients had a mixed response (at least 1 progressive lesion and 1 non-progressive lesion). This is consistent with the study from Trebeschi *et al*, which also found that combined predictions made on individual lesions was associated with OS with a significant survival difference at 1 year of 25% (77% vs 52%, log rank p = 0.02). Mixed response has been linked with poorer outcomes in several advanced cancer types [18], [48]–[51]; as such, it would not be unexpected to observe the same in our dataset. Curiously, the model for lesions receiving DM was able to achieve significant performance, especially compared to the model for lesions receiving DE. It is possible that there is an underlying biological effect which could explain lesion response to DE that imaging is simply not as sensitive to. The differential performance observed between the models for the DM and DE regimens is again consistent with Trebeschi *et al*; predictive performance for lesion level response prediction was evaluated and variation in AUROC was observed between intervention types (immunotherapy versus chemotherapy) [18].

Baseline lesion volume was significantly different between response categories, irrespective of treatment. Specifically, we observed that progressive lesions were smaller in size compared to non-progressive lesions. Evofosfamide becomes activated in hypoxic, low pH environments which have been shown to be present in sarcoma but is usually associated with larger sarcomas or sarcomas with more tumor heterogeneity related to differences in blood perfusion and/or tumor necrosis [24]. It is possible that smaller and/or more homogeneously enhancing lesions would not contain the type of environment leading to activation of evofosfamide, which may explain our observation in the DE regimen. Mathematical models for tumor growth kinetics have been widely used in precision oncology. One of the main findings from early studies is that tumor growth is not entirely exponential; over time, left unperturbed, the specific growth rate of a tumor can slow down [52]. The lesion growth in our study was perturbed in the sense that an intervention was given, hence it is unclear if our observation can be fully explained theoretically.

Given the difference in baseline volume between response categories, we removed volume-correlated radiomic features from our analysis. Previous work has shown that removal of these volume-dependent features can negatively affect model performance in the setting of patient-level risk stratification. Although removing these features may have ultimately affected our model performance, we believe that machine learning methods should have a good trade-off between transparency, performance and quality of fit. Both models included a feature calculated from a derived sub-volume of interest (the lesion core and interior rim). To this end, our results align with Sun et al, as 3 features in their validated signature are sourced from a peripheral ring [53]. While we would not expect the features to be the same as the task is different (predicting immunotherapy versus chemotherapy response), the inclusion of peritumoral features is nonetheless interesting and likely worth further exploration in future radiomics studies.

Our study has potential limitations. While our model achieved strong performance to predict lesion-specific systemic treatment response in pulmonary metastases, relying solely on volume changes at one site may not provide a comprehensive picture of the tumor’s behavior. Cancer is a complex disease that can spread to other areas of the body, and metastatic lesions may behave differently with respect to one another and the primary tumor. We chose to focus on the lung as it was the most common metastatic site in our patient population and therefore represented the most populous lesion population which was therefore less prone to produce statistical issues related to overfitting and multiple testing. The lung is also a common metastatic site in many other cancers, including but not limited to colorectal, head and neck, breast and urologic cancers [54]. Conducting prospective validation studies in LMS and other sarcomas will provide important evidence regarding the clinical utility and generalizability of your signature across different tumor types. It will help assess the performance of the model in real-world settings and validate its ability to predict lesion-level response accurately. Furthermore, monitoring multiple sites within a patient’s tumor can provide valuable insights into the heterogeneity of treatment response. Tumors can exhibit spatial variation in their response to therapy, with some sites showing a positive response while others may not. By monitoring multiple sites, it is possible to gain a more comprehensive understanding of the tumor’s overall response to treatment. We leave prospective validation, extension to other sarcomas, and monitoring of multiple sites for future work.

### Conclusion

This work introduces a radiomic-based model for predicting lesion-level response to standard systemic treatments in metastatic LMS patients, aiming to personalize treatment approaches and enhance decision-making in patient management. The model utilizes advanced machine learning techniques to tailor treatment predictions to individual lesions, exemplifying precision medicine principles in oncology. By adapting therapies based on lesion-specific characteristics, it has the potential to optimize treatment strategies and improve patient outcomes, particularly for multi-metastatic cancer patients. Continued research and validation efforts are crucial for further strengthening the evidence base and potentially translating these findings into clinical practice for the benefit of patients.

### Informed Consent and Patient Details

This study was performed in accordance with relevant guidelines and regulations. Given the retrospective and non-interventional nature of the study, a waiver of informed consent was granted. The study protocol was approved by the University Health Network (UHN) Research Ethics Board (REB), IRB no. 2020-57.

## Data Availability

The individual participant imaging data are confidential, but may be obtained with Data Use Agreements from the Sarcoma Alliance for Research through Collaboration (SARC, https://sarctrials.org/). However, the deidentified radiomic features dataset along with the code for data cleaning and analysis that underlie the results reported in the article is publicly available at https://github.com/caryn-geady/Lung-Lesion-Treatment-Response-Prediction.git and https://codeocean.com/capsule/2540686/tree/v1.

https://github.com/bhklab/Lung-Lesion-Treatment-Response-Prediction

https://codeocean.com/capsule/2540686/tree/v1

## CRediT authorship contribution statement

CG performed the analysis, generated all figures and tables and wrote the main manuscript text in consultation with SS, DB and BHK. FAB reviewed the statistical procedures. AK reviewed the image segmentations. All authors reviewed the manuscript.

## Acknowledgments

Research reported in this publication was supported by the National Cancer Institute of the National Institutes of Health under Award Number P50CA272170. The content is solely the responsibility of the authors and does not necessarily represent the official views of the National Institutes of Health. The authors would like to thank the Sarcoma Alliance for Research through Collaboration (SARC) for access to the imaging data used in this study.

## Supplementary Material

### Selected Features

**Table S1:** Summary of radiomic features included in best performing models for the prediction of pulmonary lesion-specific systemic treatment response to DM.

| Group | Radiomic Feature |  |  |
| --- | --- | --- | --- |
|  | Filter | Class | Name |
| All lesions | Wavelet-LHL | First Order | Variance (exterior rim) |
| | LOG ($\sigma = 3\text{mm}$) | GLDM | Grey Level Variance (lesion core) |
|  | - | Shape | Voxel Volume |
| \|Z\| <= 3 | Wavelet-LHL | First Order | Variance (exterior rim) |
| | LOG ($\sigma = 3\text{mm}$) | GLDM | Grey Level Variance (lesion core) |
| | LOG ($\sigma = 3\text{mm}$) | First Order | 10th Percentile (exterior rim) |
|  | - | Shape | Voxel Volume |

**Table S2:** Summary of radiomic features included in best performing models for the prediction of pulmonary lesion-specific systemic treatment response to DE.

| Group | Radiomic Feature |  |  |
| --- | --- | --- | --- |
|  | Filter | Class | Name |
| All lesions | LOG ($\sigma = 4\text{mm}$) | GLDM | Grey Level Variance (interior rim) |
|  | Logarithm | GLCM | Sum of Squares (lesion core) |
|  | - | Shape | Voxel Volume |
| $\vert Z\vert \leq 3$ | Wavelet-HHL | First Order | Variance (lesion core) |
|  | Wavelet-LLH | First Order | Mean (lesion core) |
|  | Logarithm | First Order | Mean (lesion core) |
|  | Wavelet-LHL | GLCM | Cluster Tendency (lesion core) |
|  | - | Shape | Voxel Volume |
| $\vert Z\vert > 3$ | Logarithm | GLCM | Sum of Squares (lesion core) |
|  | Logarithm | First Order | Minimum (interior rim) |
|  | Wavelet-HHL | First Order | Variance (lesion core) |
|  | Wavelet-LHL | GLCM | Cluster Tendency (lesion core) |
|  | - | Shape | Voxel Volume |

### Hyperparameter Tuning

Hyperparameter tuning was performed with GridSearchCV in Python. This process systematically explores optimal hyperparameters for machine learning models. It involves cross-validation, where the dataset is divided into folds, and each hyperparameter combination is assessed using a separate fold. GridSearchCV automates this process by exhaustively searching through a predefined grid of hyperparameter values to select the best-performing combination based on a specified evaluation metric. The predefined grid of hyperparameters used for this study are in **Table S3**.

**Table S3:** Predefined grid of hyperparameters used for GridSearchCV used in this study.

| Hyperparameter | Options |
| --- | --- |
| 'penalty' | ['l1', 'l2', 'elasticnet', None] |
| 'solver' | ['lbfgs', 'liblinear', 'newton-cg', 'newton-cholesky', 'sag', 'saga'] |
| 'tol' | [1e-4, 1e-5, 1e-6, 1e-7, 1e-8, 1e-9] |
| 'max_iter' | [50,100,150,200] |

